# Prevalence of use of mobile food delivery services during school hours in the United States: Survey study

**DOI:** 10.1101/2025.11.12.25339631

**Authors:** Mika Matsuzaki, Audrey W Ting, Nick Birk, Maria E Acosta, Carla Tarazona-Meza, Brisa N. Sanchez, Emma Sanchez-Vaznaugh, Sabri Bromage

**Affiliations:** Johns Hopkins Bloomberg School of Public Health, Department of International Health; Mahidol University, Institute of Nutrition; Harvard T.H. Chan School of Public Health, Department of Biostatistics; San Francisco State University, Department of Public Health; Drexel University Dornsife School of Public Health, Department of Epidemiology and Biostatistics; Harvard T.H. Chan School of Public Health, Department of Nutrition

## Abstract

**Background:** Federal and state school nutrition policies over the past 20 years have improved the nutritional quality of school meals, children’s diet quality, and childhood obesity prevalence in the United States. However, increasing use of mobile food delivery apps during school hours may introduce new dietary risks among adolescents.

**Objectives:** This study aimed to assess the adolescent usage patterns of and perceptions toward mobile food delivery during school hours in the United States.

**Methods:** We administered a national online survey in July-August 2025 using the AmeriSpeak Teen Omnibus platform administered by the National Opinion Research Center at the University of Chicago. The respondents were 1,027 adolescents aged 13-17 years. We estimated and statistically compared survey-weighted distributions of self-reported types and frequencies of mobile food app usage characteristics during and after school by age, sex, race/ethnicity, household income levels, and regional strata using Chi-squared tests. We also conducted thematic analyses of responses to an open-ended question asking why participants supported or opposed the usage of mobile food delivery services during school hours.

**Results:** Nearly 1 in 4 adolescents used mobile food delivery services during school hours and nearly half used them after school. Large proportions of adolescents (58.8% and 34.1%) used these services to order fast foods and sugar-sweetened beverages, respectively, while grain bowls, fruit, non-deep fried vegetables, and unsweetened beverages were less popular. About 34% of adolescents in the western U.S. attended schools that allowed mobile food delivery during school hours, a substantially higher proportion than other national regions; however, adolescents in the west more frequently cited the perceived high costs as the reason for not using those services. Nearly half of the respondents (47.2%) support the idea of mobile food delivery during school hours. However, distraction from their learning environment was a major concern regardless of their support or opposition to mobile food delivery at school.

**Conclusions:** Mobile food delivery during school hours is a relatively new method of food acquisition for adolescent students. School nutrition policy should consider students’ access to and usage patterns of both physical and digital food environments to help ensure development of lifelong healthy eating habits among adolescents.

## Introduction

Diet and nutrition are key determinants of childhood obesity [1,2]. Studies over the past two decades have shown that unhealthy food environments are risk factors for poor dietary behaviors and obesity among youth. [3,4]. Access to food from outside of schools was historically possible only in schools where students were allowed to leave campus during lunchtime. In recent years, in addition to physical food environment risk factors like the proximity to unhealthy food outlets, *digital food environments*, such as online food delivery, have shaped youth dietary choices [5,6]. Online food delivery enables youth to obtain outside foods in lieu of or in addition to foods served in schools. However, the usage pattern of and perceptions towards online food delivery platforms during school hours has not been examined.

The emergence of digital tools for food acquisition has introduced additional challenges in measuring and enabling healthy food environments [7]. Digital food environments influence dietary behaviors through various platforms for online food shopping, and also food and health information on the web, social media, and online advertisements [5,6]. These platforms can influence food choices and dietary patterns in tandem with physical food environments. Although agencies like the World Health Organization Regional Office for Europe have raised concerns regarding the impact of digital food environments, especially for youth, the efforts to assess and develop strategies to promote healthy dietary behaviors with consideration to the combination of physical and digital food environments remain underdeveloped and fragmented [5].

In the United States, local, state, and federal policies have been enacted to improve the food environment inside schools such as the federal Healthy, Hunger-Free Kids Act 2010 and SB12/965 in California, covering a wide range of topics from banning sugar-sweetened beverages and requiring whole grain inclusion to setting nutrition standards of school meals like sodium, calories from saturated fat [8–10].

Studies have shown evidence of beneficial effects of these regulations of school food environments on students’ diets and nutritional status [11–16]. However, policies and policy evaluations have not incorporated the recent surge in the use of food delivery apps among youth. If a large number of schoolchildren order food from outside school to substitute consumption of school meals, the impact of school nutrition policies would be diminished. In the 2025-2026 academic year, 31 states ban or restrict the use of mobile phones altogether at school, which would prevent students from making mobile food delivery orders [17,18]. However, the extent to which schools have applied additional rules specifically for mobile food delivery (e.g., explicit bans or exceptions to the rules) remains unclear. Furthermore, even when general use of mobile phones is banned or restricted during instruction periods, phone use during lunchtime may be allowed, or mobile food delivery may be facilitated via schoolteachers and other staff, before or after school hours, or otherwise against rules. However, the prevalence of mobile food delivery during school hours in the United States is currently uncharacterized, presenting a significant gap in our understanding of how digital food environments influence student dietary behaviors.

This study aimed to address this gap through a US national survey among school-going adolescents. Using the National Opinion Research Center’s AmeriSpeak Teen Panel platform, we conducted an online survey with over one thousand adolescents to assess and analyze 1) usage patterns of food delivery apps during and after school hours and 2) adolescents’ perspectives on whether usage of these apps to get food delivered from external food outlets should be allowed in schools.

## Methods

### Sample

We conducted an online survey (July 31-August 13, 2025) among a national sample of 1,027 English-speaking, U.S. adolescents aged 13-17 years old through the National Opinion Research Center (NORC) AmeriSpeak Teen Panel Q3 Teen Omnibus survey platform [19]. AmeriSpeak surveys are designed to be nationally-representative of the U.S. household population by randomly sampling from the NORC’s National Frame and complementary databases [20]. The AmeriSpeak Teen panel, constructed by recruiting teens from AmeriSpeak households with children aged 13-17 years old, has a recruitment rate of 26.3% and a retention rate of 78.0%. The AmeriSpeak panel participants were recruited by NORC using several methods including mail, phone, and in-person field interviews in English and Spanish [20]. After consent and an initial survey, registered panelists were invited to participate in web- or phone-based research studies. In this study, our survey questions were released to 1,444 AmeriSpeak Teen panelists, and 723 completed the survey. NORC aimed to provide a minimum of 1000 respondents and recruited additional teens from a non-probability-based, opt-in sample (n=304). The two samples were then combined using NORC’s TrueNorth 3.0 calibration tool, a tree-based non-parametric supervised learning algorithm that creates combined sample weights for sample non-coverage and bias correction. TrueNorth 3.0 has been proven to reduce bias by more than half compared to nonprobability sample estimates [21]. A raking process was used to adjust for any survey nonresponse as well as any noncoverage or under- and oversampling in both probability and non-probability samples resulting from the study-specific sample design. Raking variables for both the probability and nonprobability samples included age, sex, census region, race/ethnicity, and parent’s highest education. Population control totals for the raking variables were obtained from the February 2024 Current Population Survey. The weighted data reflect the U.S. population of teens aged 13 to 17. The recruitment, survey administration, and data cleaning were done by the NORC team. The research team received only the anonymized data from NORC. The detailed procedure of the NORC AmeriSpeak Panel Surveys is described elsewhere [19].

#### Ethical Considerations

The Institutional Review Board of the Johns Hopkins Bloomberg School of Public Health determined this study to be exempt from the human subject research regulations, as the team only received data from NORC in which participants cannot be re-identified (IRB00033369). The Institutional Review Board at NORC (Federal Wide Assurance #FWA00000142) reviews research protocols before any NORC survey begins. [22]. Informed consent for AmeriSpeak panel participants was obtained either online or by phone [20]. The participants receive point-based compensation for participating in the AmeriSpeak survey ($1 per 1000 points), which are redeemed when a minimum of $10 is achieved. For this survey, they received 4000 points.

### Measures

To assess food delivery app usage patterns, the survey included questions regarding: frequency of current use of mobile apps to order food for delivery to school during and after school hours, the length of time since the school has allowed mobile food delivery apps (if any) was enacted (≥5 years or <5 years), types of foods typically ordered on these apps, and perspectives on whether and why schools should allow/disallow use of these apps. The survey questions were reviewed and tested by three experts on the team for content and face validity and for clarity of the wording. The survey was iteratively tested and improved until all three researchers were satisfied. Because this survey was administered as part of the routine survey that is administered by NORC, we did not pilot the survey in a small sample of the target population. We specifically asked for the usage pattern during school hours, as the survey was conducted in the summer months. Mobile food app delivery usage frequency was categorized as frequent (at least once a week), infrequent (less than weekly), or never. For the choice “Never”, the respondents also chose one of three reasons: the school does not allow such usage, it is too expensive, or other non-specified reasons. We also asked whether the respondents favored, opposed, or were unsure about whether mobile food delivery should be allowed during school hours. This question was followed by an open-ended question probing the reasons why they chose those responses. NORC AmeriSpeak provided data on participant sociodemographic and economic backgrounds (adolescent-reported age, sex, and race/ethnicity; and parental-reported household geographic location, highest education levels, and household income), types of devices used to answer the survey, and survey weights. NORC AmeriSpeak provided the four household income categories included in the analysis.

### Analysis

Statistical analyses were performed in R v4.5.1. The current study focused on descriptive analyses of the survey responses, including survey-weighted percentages and unweighted frequencies. Unweighted percentages based on the total sample and on the AmeriSpeak Panel sample are shown in Supplemental Table 1. Weighted distributions of reported usage of food delivery apps during and after school were stratified by sociodemographic, economic, and geographic characteristics, and statistically compared using Chi-squared tests. For each survey question, we removed missing data points. Weighted statistics and hypothesis tests were conducted using the “survey” package in R.

For the open-ended questions, we followed a recursive 6-step process: Familiarization of the data, generation of initial codes, theme construction, theme reviews, naming of themes, and reporting of the results [23]. A total of 814 adolescents answered the open-ended question on perceptions of school policy allowing food delivery apps. Some responses were assigned to more than 1 code. One author assigned codes to all responses first using Taguette software. the assignments were checked by two other authors, who assigned additional codes, which in turn were reviewed by the first author. Two authors conducted an inductive thematic analysis of 10% of the responses to develop the themes. The primary purpose of team coding and analysis was to ensure robustness and clarity of our framework. We resolved all coding disagreements through iterative discussions among the coding team until a consensus was reached. We then grouped similar codes into themes. We iteratively refined the themes throughout this process. The initial set of 183 sub-themes was grouped into 26 themes (Supplemental Material 3), excluding 10 tags assigned to the responses that were unclear or not applicable (e.g., “homeschooled”). We analyzed the frequencies of the themes from the thematic analyses to assess the most common reasons for supporting or opposing policy restricting the use of mobile apps for ordering food during school hours. To ensure the credibility and confirmability of the findings, the inductive thematic analysis was grounded strictly in the participants’ textual responses. The analysis aimed to reflect the explicit meanings communicated by the respondents. We maintained records of iterative code and theme development and multiple team members were involved in the analysis. The responses were from a diverse, national sample, which helps capture a broad spectrum of experiences across regions and sociodemographic contexts. This exploratory analysis aimed to offer the first insights into how students perceive the option of mobile food delivery during school hours.

While all authors have conducted nutrition research in U.S. settings, we currently do not work in or attend high schools, limiting direct knowledge of current high school students’ day-to-day experiences. Two researchers interact with secondary schools as parents. Four researchers regularly conduct school nutrition research. One researcher worked in secondary schools before working as a researcher. These past interactions with students, school systems, and/or nutrition data on adolescents may have influenced our development and interpretations of the themes. All authors are familiar with mobile food delivery services.

## Results

This national survey of adolescents involved 1027 respondents, 49.1% female, with a mean age of 15.2 ±1.3 years. A majority of respondents (76.7%) used smartphones to complete the survey. A quarter of the adolescents were currently using—either frequently or infrequently—mobile food delivery services during school hours (Table 1). The majority of students (72.0%) said that their schools never allowed mobile food delivery during school hours while 7.5% reported that their schools had allowed this practice for more than 5 years and 18.5% less than 5 years. The use of these services was more common after school (49%). Chi-squared tests for differences in usage of mobile food delivery apps during school hours showed differences by race/ethnicity, household income, and region. About one-fifth of Black adolescents frequently used these services during school hours, while smaller proportions of the adolescents from other racial/ethnic groups reported frequent usage. Twenty-nine percent of the adolescents from the lower-income groups for households (≤$30,000 and $30,000 to ≤60,000) used these services during school hours in comparison to 23% of adolescents from higher-income households. This response difference by household income levels was not seen for after-school usage.

**Table 1.**
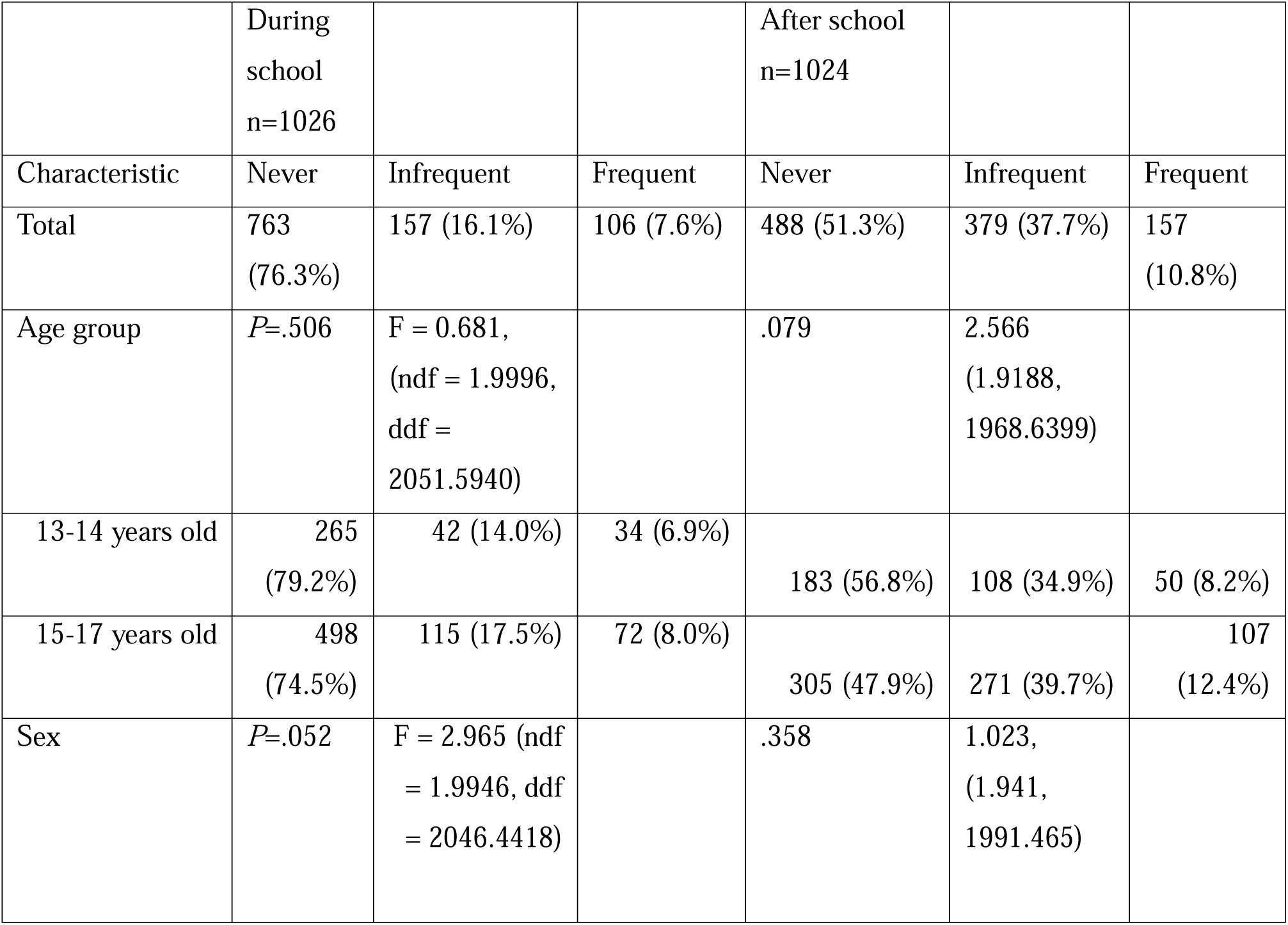
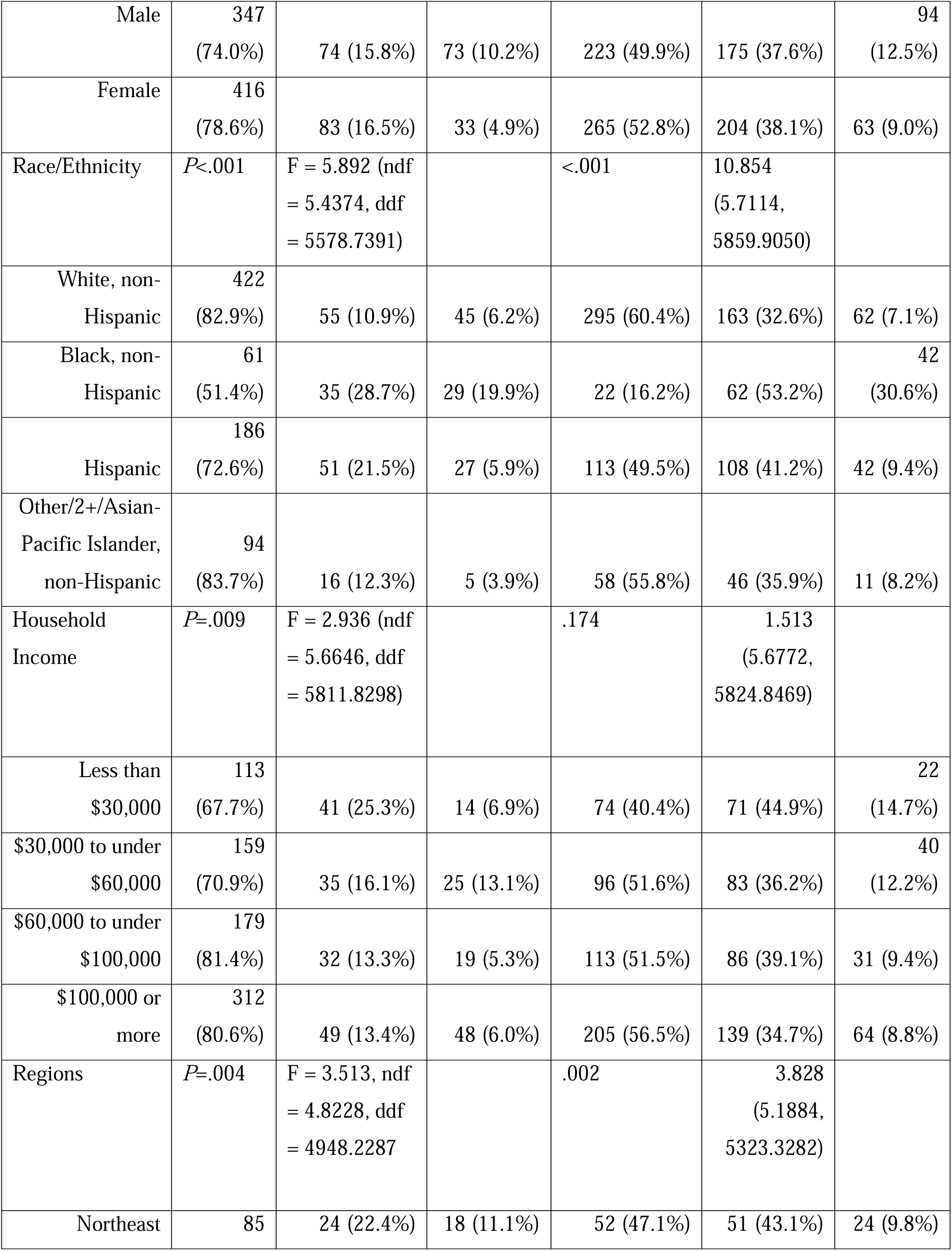
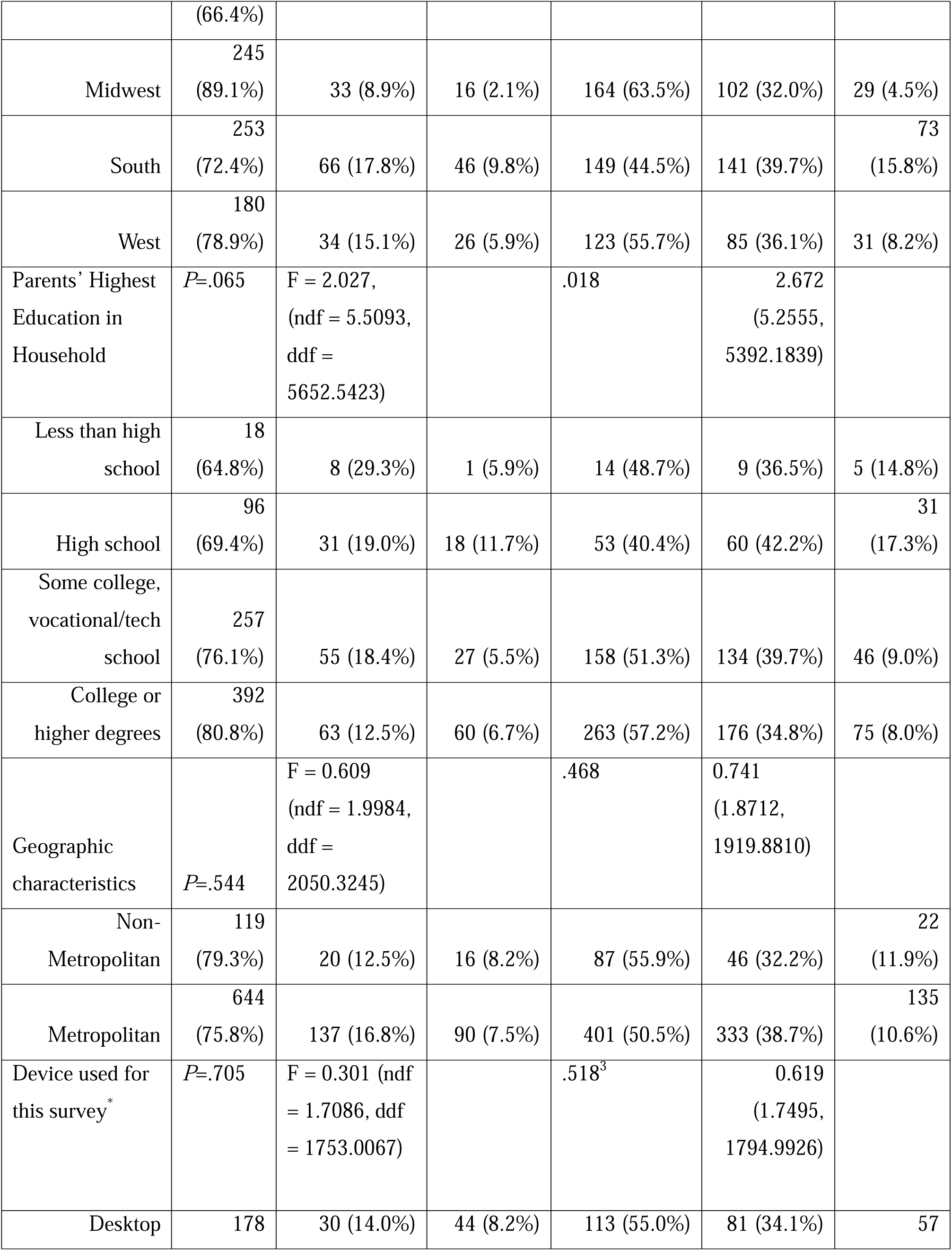
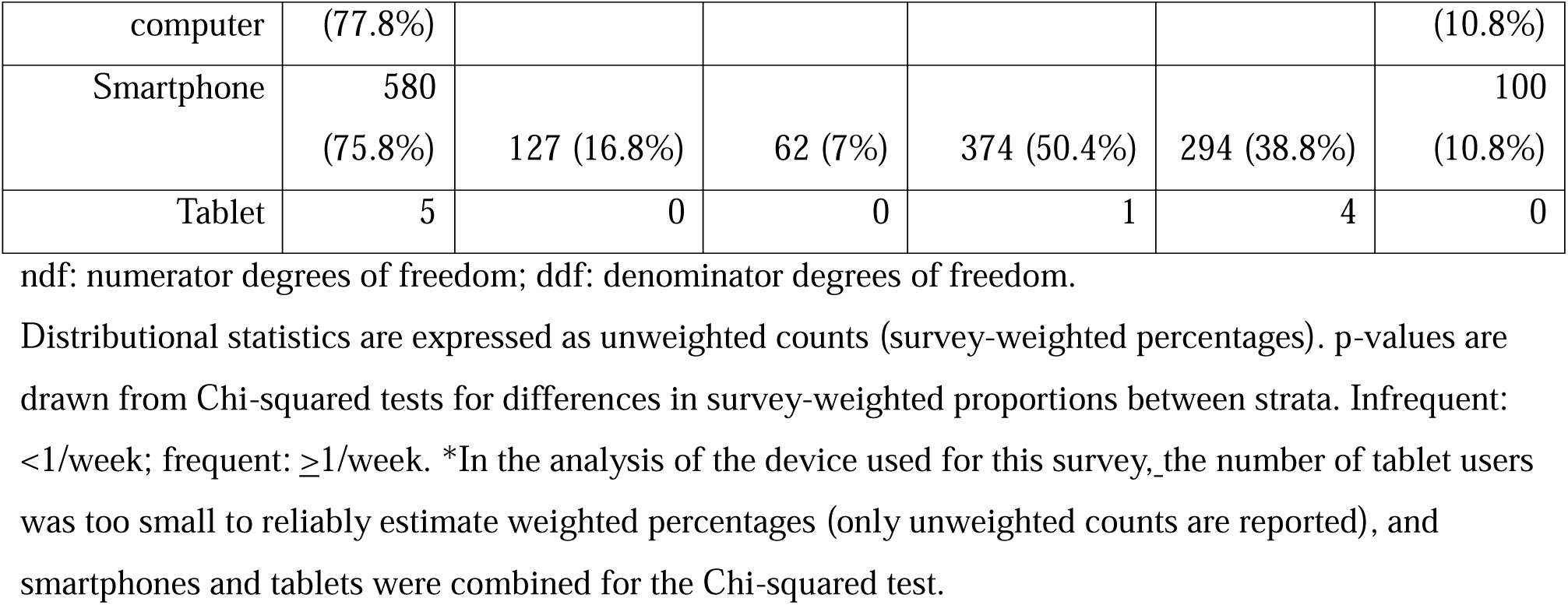
Frequency distribution of mobile food delivery app usage during and after school by participant and household characteristics.

The proportion of adolescents who responded that their schools allowed mobile food delivery was highest among those from the West (34.5%), compared to 22.1%, 24.4%, and 24,9% in the Northeast, Midwest, and South regions, respectively (Supplemental Table 2). However, while a greater proportion of schools in the West allowed these services during school hours, adolescents in the West more commonly cited the perceived high costs as the reasons for not using these services than the other regions (Supplemental Table 3 and 4).

Participants were asked what they ordered in mobile food delivery apps, whether at school or elsewhere. The overall patterns of types of foods ordered online were similar across racial/ethnic groups (Figure 1), with fast food items (burgers, burritos, tacos, or pizza) reported commonly across subgroups and orders for grain bowls, non-fried vegetables, and fruits less frequently reported. For beverages, 34.1% of adolescents ordered sugar-sweetened beverages (SSBs) in comparison to 5.3% for unsweetened beverages and 16.1% for beverages with artificial sweeteners. The analysis of all answer types—including combinations of these multiple choices—showed fast food or fast food in combination with SSB as the two most common online orders.

**Figure 1.**
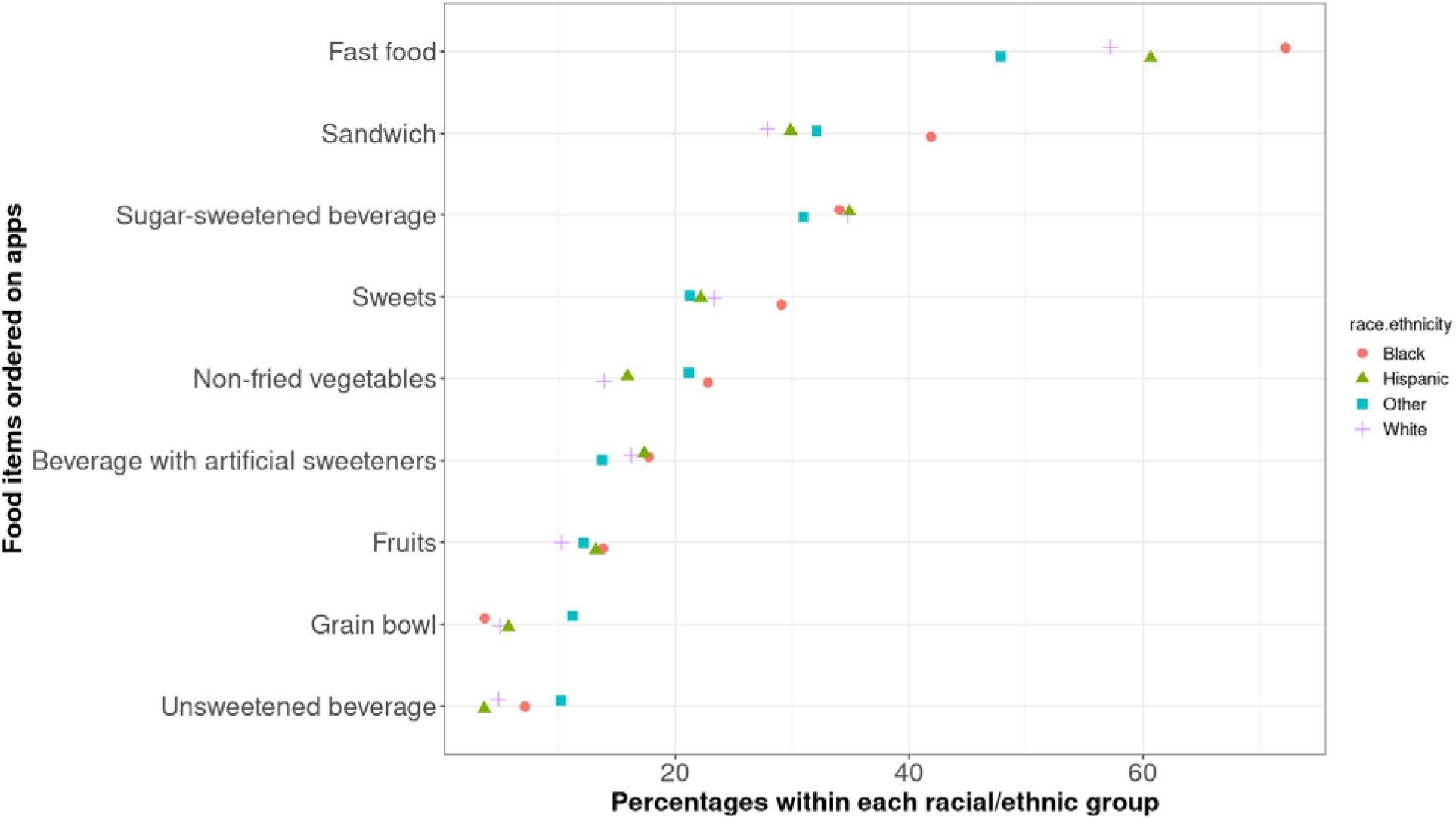
**Types of items ordered via food delivery apps by the adolescents by race/ethnicity in 2025. Multiple choice selection was allowed in this question. The values are weighted percentages.**

While nearly half of the respondents responded affirmatively when they were asked whether they should be allowed to use mobile apps for food delivery during school hours, the other half were either unsure or disapproving (Table 2). We did not find differences in response patterns for any of the subgroups.

**Table 2.** Distribution of responses to the question on whether participants supported the usage of mobile food delivery apps during school hours (n=1018)*

|  |  | Yes<br>% | No<br>% | I'm not sure<br>% | *F* (*ndf*, *ddf*) | *P*-value<sup>1</sup> |
| --- | --- | --- | --- | --- | --- | --- |
|  | Total | 47.4 | 28.7 | 23.9 |  |  |
| Age | 13-14 | 43.4 | 30.3 | 26.3 | 1.062<br>(1.9944,<br>2028.3088) | .346 |
|  | 15-17 | 49.9 | 27.8 | 22.4 |  |  |
| Sex | Female | 47.6 | 30.2 | 22.3 | 0.476<br>(1.9983,<br>2032.2232) | .621 |
|  | Male | 47.3 | 27.3 | 25.4 |  |  |
| Highest parental education in household | Less than high school | 66.7 | 17.6 | 15.6 | 0.742,<br>(5.4908,<br>5584.1012) | .604 |
|  | High school | 44.9 | 28.5 | 26.5 |  |  |
|  | Some college or vocation/tech schools | 49.0 | 26.6 | 24.4 |  |  |
|  | College/higher degrees | 46.4 | 30.7 | 22.9 |  |  |
| Race/ethnicity | White, Non-hispanic | 41.7 | 33.8 | 24.5 | 1.869<br>(5.6184,<br>5713.9623) | .09 |
|  | Black, Non-Hispanic | 59.5 | 19.8 | 20.6 |  |  |
|  | Hispanic | 51.9 | 26.3 | 21.7 |  |  |
|  | Other/Multiple, Non-Hispanic | 48.3 | 22.0 | 29.8 |  |  |
| Household income | <\$30k | 53.5 | 23.3 | 23.2 | 1.246<br>(5.8416,<br>5940.9361) | .280 |
| | \$30k-<\$60k | 52.2 | 26.5 | 21.4 | | |
| | \$60k-<\$100k | 46.4 | 26.0 | 27.5 | | |
| | \$100k+ | 42.1 | 34.5 | 23.3 | | |
| Region | Northeast | 38.6 | 35.9 | 25.5 | 0.919,<br>(5.5922,<br>5687.2948) | .475 |
|  | Midwest | 49.7 | 27.4 | 22.8 |  |  |
|  | South | 47.0 | 27.2 | 25.8 |  |  |
|  | West | 52.4 | 27.4 | 20.2 |  |  |

ndf: numerator degrees of freedom; ddf: denominator degrees of freedom.
Distributional statistics are expressed as unweighted counts (survey-weighted frequencies \* We removed 9 respondents who skipped this question from the analysis. 1. p-values are from drawn the Chi-squared tests for differences in survey-weighted proportions between strata

The analyses of the open-ended questions asking why or why not the respondents believed those services should be allowed in school showed several common themes (Figure 2). The most common reason for supporting mobile food delivery during school hours was the desire or need for alternative options. This included preferences, desire for more diverse food selection, unmet needs for allergy or dietary restrictions, and religious reasons. The participants also commonly believed that the food at school was low quality, unhealthy, or not nutritious. Several participants reported not eating enough or at all during school hours. Time constraints during lunch hours (e.g., the lines are too long) were also a commonly reported logistical problem. The participants who were supportive of the use of mobile food delivery during school hours perceived these services to be more convenient, easy, and/or fast. Many also stated that students have or should have the right and freedom to order what they like with their own money. A few respondents also stated that this is a learning opportunity for making smart choices with online food delivery.

> “It gives us kids a sense of responsibility of growing up and manage our finances” (male respondent)

**Figure 2.**
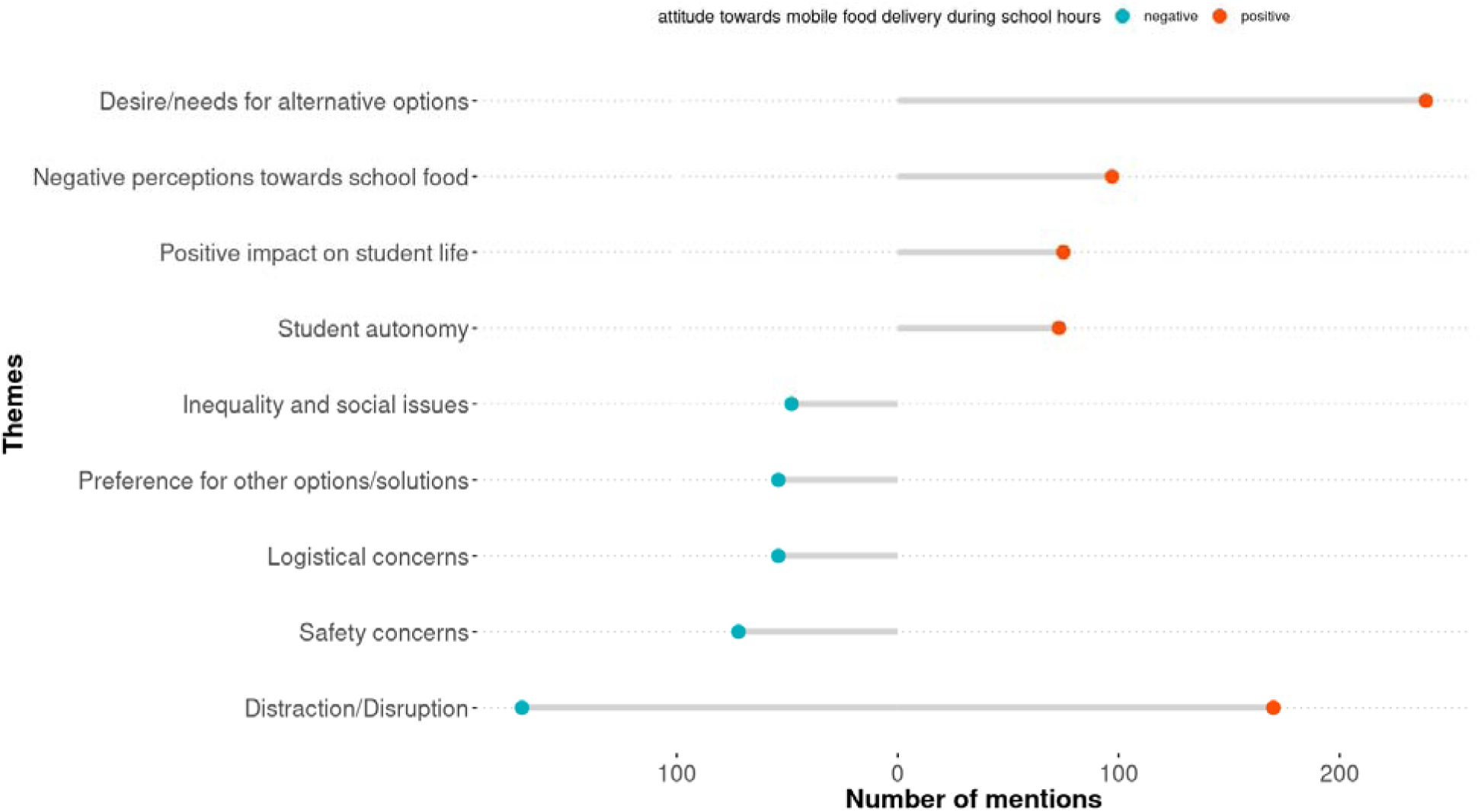
Themes commonly discussed by the survey respondents regarding whether the use of mobile food delivery should be allowed in schools. The values are the number of mentions among 814 respondents who provided relevant answers to this open-ended question. Some respondents discussed multiple themes or both negative and positive viewpoints in their individual responses.

*“It doesn’t *seem* to be a problem, not at my school at least…with the option of ordering lunches from apps, it will teach soon-to-be adults money management. If it wasn’t allowed, then they wouldn’t have the opportunity to test and develop their money skills. In the ‘real world’, no rules will prevent people from spending their money, so it’s important to give young people ‘trial runs’” (female respondent)*

On the other hand, many respondents who were conditionally supportive or unsupportive of mobile food delivery in school were concerned that allowing these services would introduce distractions and disruptions to the school environment. The participants were also wary of unfairness, with only those who can afford to purchase food online would do so, and highlighted potential concerns about stigma and bullying. Safety concerns were also common, with many respondents discussing random visitors coming to school and potentially causing problems. Even respondents who were conditionally supportive of mobile food delivery at school mentioned these potential issues:

> “I think students should be allowed to order food on apps at school, as long as it’s done responsibly. It gives them more choice, can accommodate dietary needs, and is helpful when cafeteria options are limited. That said, schools should set clear rules to avoid distractions or safety issues” (male respondent)

> “It depends because if done well and orchestrated it works but that would require well behaved kids or motivated staff, or maybe both” (male respondent)

## Discussion

To our knowledge, this is the first study to show the prevalence and perceptions of mobile food delivery in school, based on a national sample in the U.S. We found that 1 in 4 adolescents are using mobile food delivery during school hours, and about half of the adolescents use these services after school. Fast food items and sugar-sweetened beverages were the most popular online orders among these adolescents. Our results suggest that schools in the West may be allowing the use of online food delivery platforms more but more adolescents in the West may choose not to use these services because of the perceived high costs. About half of the respondents supported the idea of schools allowing the usage of online food delivery during school hours, although many also noted concerns like distractions to the learning environment and logistic complications.

Online food delivery has become exponentially popular globally over the past decade. While a five-country survey reported 15% of the respondents using online food delivery in 2018—11% in the U.S.[24], the COVID-19 pandemic accelerated this trend. Post pandemic, the prevalence of online food delivery usage quadrupled for full-service restaurant online orders in comparison to the pre-pandemic period [25]. As seen in our survey responses, regional differences in online food delivery use and perceptions may exist—in 2025, DoorDash reported the highest number of complete orders in the West [26]. Although research on the impact of increased use of online food delivery services on health outcomes is still in its nascent stage, studies have long suggested adverse effects of eating food from outside of home, such as increased energy intake from fat and lower intake of micronutrients [27]. Among children, higher consumption of fast food is associated with intake of greater total calories, total fat, total carbohydrates, added sugar, sugar-sweetened beverages, and less fiber, fruits, and non-starchy vegetables, all of which are risk factors for obesity and nutrition-related chronic diseases [28]. Among our study participants, unhealthy foods and drinks were common online orders, similar to the findings on youth from other countries [29,30]. The prior work points to potential negative effects of online food delivery on youth’s diet and health [31].

Over the past decade, news media outlets have reported on adolescent use of food delivery apps in schools and schools’ reactions [32], but no systematic study examined the extent of usage in schools. The current study found that nearly 1 in 4 adolescents are using these services in school. Some adolescents mentioned that their schools have a blanket policy banning or restricting the use of mobile phones during school hours and preventing them from ordering food online, which is consistent with the reports of state-level policies on restrictions on phone use during school hours [17]. There are currently no federal policies on whether or how online food orders should be regulated in schools.

Studies have found a positive impact of the federal and state school nutrition policies on student diet and weight status [11,12,33]. However, these policies generally only extend to the food environment within schools, but students acquire food during school hours from other sources like food outlets near schools and, more recently, mobile food delivery. Several studies have found evidence of positive associations between physical food environments near schools and body weight or weight status of schoolchildren [34–36]. While it is unclear whether and how much food environments may modify the effects of school nutrition policies, one study found that students attending schools with closed campus policies consumed fast food places or restaurants less frequently, offering insights on the potential impact of outside foods delivered via online platforms [37]. Our study found that fast food items were popular online orders among these adolescents and potentially healthier items like fruits and grain bowls were less commonly reported, which may lead to poor diet among adolescents attending schools that allow online food delivery.

### Implications

Given the pervasiveness of online food orders in society, schools and policymakers should discuss how best to protect youth from the harmful effects of digital food environments, both in terms of the short- and long-term impact. A ban on mobile food delivery alone may not teach adolescents how to navigate through digital food environments and make smart choices in the modern physical and digital food environments during and beyond schooling years. Secondary schools offer an excellent venue to provide such education. Digital literacy has long been recognized as an important part of modern education with Information and Communication Technology, and digital literacy programs have been incorporated into the curriculum in many school globally [38]. However, digital literacy training programs largely focus on how to use digital tools for their learning and work. There are missed opportunities to teach youth through school curriculm how to interact with digital food environments, from online food purchases to nutrition information on social media. This could also be part of the updated health education requirements to include these competencies within the Education Code, as most states already specify core health concepts that must be taught [39]. Future research can assess the effects of the intertwined physical and digital food environments on youth dietary behaviors in observational and multi-component intervention studies. There is a need for a different conceptual framework to think about digital food environments, as these digital tools may change the concept of proximity (e.g., food delivery from places that are not near schools). Online food shopping is recognized as an important alternative means of food acquisitions in other national context such as for the users of Supplemental Nutrition Assistance Program in the U.S., with an increasing awareness of the needs to education the users how to use these digital tools [40–42].

Collaborative partnerships between researchers, governmental stakeholders, and online retailers may be able to help develop intervention programs, curriculum, and policies that help adolescents develop skills and knowledge needed for life-long healthy dietary behaviors. Partnerships with online platforms with strategies like discounts and more prominent displays of healthier food items may also help youth make smart food choices online.

### Strengths and limitations

The current survey was the first to use a national sample to estimate the prevalence of mobile food delivery usage among adolescents in the U.S., with sociodemographic and regional information allowing us to examine responses by subgroup. Our results also included open-ended questions to which adolescents explained why they support or do not support the use of these services at school, providing deeper insights into their perspectives around online food delivery and, more generally, diet during school hours. On the other hand, the prevalence of service usage and school policies were self-reported data and under or overestimation may have occurred. While we cannot confirm whether these reports reflect the reality, we included a question to gauge confidence in their knowledge on school policies and checked their answers to multiple-choice questions against their responses to the open-ended question for consistency. Our survey was administered during the summer break, which may have introduced recall bias on behaviors during the academic year. We did not perform psychometric validation of the instrument as our questions were included in the pre-scheduled, quarterly survey of the AmeriSpeak Panel. However, our survey was descriptive in nature and our iterative development of the survey with the expert reviews provided a set of sound exploratory questions, providing a starting point for further improvements in the instrument and confirmatory research. Future national studies with the schools and actual usage data—potentially from the online food delivery companies—can verify the findings from the current study and further examine geographical variabilities.

## Conclusion

Food environments surrounding adolescents have changed dramatically over the past decade. We found that about a quarter of the adolescents may be using online food delivery while in school. Adolescents were more prone to ordering fast food and sugar-sweetened beverages, and this may possibly be due to their affordable prices, speed, and preferences for taste. In addition to restrictions on mobile food delivery in school, schools should consider offering education on digital food environments and/or subsidies and other incentives for healthier foods, in partnership with the online food delivery companies, to help students make smart food choices online within and outside of school settings. Future research can further strengthen the evidence base by assessing the usage patterns of mobile food delivery among students with direct measures (e.g., receipts) and the presence of school policies from school websites and surveys.

## Data Availability

All data produced in the present study are available upon reasonable request to the authors

## Disclosure

**Author Contributions:**

MM: Conceptualization, Data curation, Formal analysis, Funding Acquisition, Investigation, Methodology, Project administration, Resources, Supervision, Visualization, Writing—original draft, Writing—review&editing.

AWT: Data curation, Formal analysis, Investigation, Project administration, Writing—review&editing.

NB: Formal analysis, Methodology, Writing—review&editing.

MEA: Conceptualization, Writing—review&editing.

CTM: Formal analysis, Writing—review&editing.

BNS: Methodology, Writing—review&editing.

ESV: Methodology, Writing—review&editing.

SB: Methodology, Writing—review&editing.

## Data availability

The datasets generated or analyzed during this study are available from the corresponding author on reasonable request.

## Use of generative AI

We used Grammarly to help with proofreading of the manuscript.

## Abbreviations

AI: Artificial Intelligence
COVID-19: Coronavirus disease of 2019
NORC: National Opinion Research Center
SSB: Sugar-sweetened beverages
U.S.: United States of America

