## Supplemental Material 1 and 2 for "Prevalence of use of mobile food delivery services during school hours in the United States: Survey study"

Supplemental Table 1: Unweighted frequencies

|  | During school n= |  |  | After school  n= |  |  |
| --- | --- | --- | --- | --- | --- | --- |
| Characteristic | Never | Infrequent | Frequent | Never | Infrequent | Frequent |
| Total |  |  |  |  |  |  |
| Age group |  |  |  |  |  |  |
| 13-14 years old |  |  |  |  |  |  |
| 15-17 years old |  |  |  |  |  |  |
| Sex |  |  |  |  |  |  |
| Male |  |  |  |  |  |  |
| Female |  |  |  |  |  |  |
| Race/Ethnicity |  |  |  |  |  |  |
| White, non-Hispanic |  |  |  |  |  |  |
| Black, non-Hispanic |  |  |  |  |  |  |
| Hispanic |  |  |  |  |  |  |
| Other/2+/Asian-Pacific Islander, non-Hispanic |  |  |  |  |  |  |
| Household Income |  |  |  |  |  |  |
| Less than $30,000 |  |  |  |  |  |  |
| $30,000 to under $60,000 |  |  |  |  |  |  |
| $60,000 to under $100,000 |  |  |  |  |  |  |
| $100,000 or more |  |  |  |  |  |  |
| Parents’ Highest Education in Household |  |  |  |  |  |  |
| Less than high school |  |  |  |  |  |  |
| High school |  |  |  |  |  |  |
| Some college, vocational/tech school |  |  |  |  |  |  |
| College or higher degrees |  |  |  |  |  |  |
| Regions |  |  |  |  |  |  |
| Northeast |  |  |  |  |  |  |
| Midwest |  |  |  |  |  |  |
| South |  |  |  |  |  |  |
| West |  |  |  |  |  |  |
| Geographic characteristics |  |  |  |  |  |  |
| Non-Metropolitan |  |  |  |  |  |  |
| Metropolitan |  |  |  |  |  |  |
| Device used for this survey^*^ |  |  |  |  |  |  |
| Desktop computer |  |  |  |  |  |  |
| Smartphone |  |  |  |  |  |  |
| Tablet |  |  |  |  |  |  |

Supplemental Table 2: Unweighted

|  | During school n= |  |  | After school  n= |  |  |
| --- | --- | --- | --- | --- | --- | --- |
| Characteristic | Never | Infrequent | Frequent | Never | Infrequent | Frequent |
| Total |  |  |  |  |  |  |
| Age group |  |  |  |  |  |  |
| 13-14 years old |  |  |  |  |  |  |
| 15-17 years old |  |  |  |  |  |  |
| Sex |  |  |  |  |  |  |
| Male |  |  |  |  |  |  |
| Female |  |  |  |  |  |  |
| Race/Ethnicity |  |  |  |  |  |  |
| White, non-Hispanic |  |  |  |  |  |  |
| Black, non-Hispanic |  |  |  |  |  |  |
| Hispanic |  |  |  |  |  |  |
| Other/2+/Asian-Pacific Islander, non-Hispanic |  |  |  |  |  |  |
| Household Income |  |  |  |  |  |  |
| Less than $30,000 |  |  |  |  |  |  |
| $30,000 to under $60,000 |  |  |  |  |  |  |
| $60,000 to under $100,000 |  |  |  |  |  |  |
| $100,000 or more |  |  |  |  |  |  |
| Regions |  |  |  |  |  |  |
| Northeast |  |  |  |  |  |  |
| Midwest |  |  |  |  |  |  |
| South |  |  |  |  |  |  |
| West |  |  |  |  |  |  |
| Parents’ Highest Education in Household |  |  |  |  |  |  |
| Less than high school |  |  |  |  |  |  |
| High school |  |  |  |  |  |  |
| Some college, vocational/tech school |  |  |  |  |  |  |
| College or higher degrees |  |  |  |  |  |  |
| Geographic characteristics |  |  |  |  |  |  |
| Non-Metropolitan |  |  |  |  |  |  |
| Metropolitan |  |  |  |  |  |  |
| Device used for this survey^*^ |  |  |  |  |  |  |
| Desktop computer |  |  |  |  |  |  |
| Smartphone |  |  |  |  |  |  |
| Tablet |  |  |  |  |  |  |
